# Updating mortality risk estimation in intensive care units from high-dimensional electronic health records with incomplete data

**DOI:** 10.1101/2022.04.28.22274405

**Authors:** Bertrand Bouvarel, Fabrice Carrat, Nathanael Lapidus

## Abstract

**Context:** Intensive care units (ICU) are subject to a high mortality rate, currently addressed by the implementation of scores (SAPS II, SOFA, APACHE II) assessing the risk of in-hospital mortality from admission data. Their performances are satisfactory to predict death when complications occur early after admission; however, they may become irrelevant in the case of long hospital stays.

**Methods:** Using the MIMIC-III database, we developed predictive models of short-term mortality in ICU from longitudinal data collected throughout patients’ stays of at least 48 hours. Several statistical learning approaches were compared, including deep neural networks and penalized regression. Missing data were handled using either complete case analysis or multiple imputation. Models’ performances were evaluated via repeated 5-fold cross-validation.

**Results:** Predictions relying on longitudinal data were more accurate than those relying solely on admission data. Complete case analyses from 19 predictors showed good discrimination (area under the ROC curve [AUC] > 0.77 for several statistical learning approaches) to predict death between 12 and 24 hours onwards, while keeping only 25% of patients in the sample. Multiple imputation allowed to include 70 predictors and keep 95% of patients, with similar performances, hence allowing predictions in patients with incomplete data. Calibration was satisfactory for all models.

**Discussion:** This proof of concept supports that automated analysis of electronic health records can be of great interest throughout patients’ stays, as a surveillance tool likely to detect lethal complications in ICU soon enough to take corrective measures. Though this framework relies on a large set of predictors, it is robust to data imputation and may be effective early after admission, as data is still scarce.

## 1. Introduction

Intensive care units (ICU) admit critically-ill patients, who require constant care and supervision from life support equipment and medication in order to ensure normal bodily functions [1]. The illness severity of patients explains the high fatality rate in ICU that remains around 20% globally [2]. Another explanation of this very high mortality rate lies in the rapid evolution of patients’ conditions and the risk of delayed management of complications. Thus, timely diagnosis and relevant management and treatment are crucial to amend prognosis. To address this issue and identify patients with the highest risks of severe complications, prognostic scores have been developed, such as the Acute Physiology And Chronic Health Evaluation II (APACHE II) [3], the Simplified Acute Physiology Score (SAPSII) [4] or the Sequential Organ Failure Assessment Score (SOFA) [5], used to predict in-hospital mortality from data collected upon admission or in the first 24 hours in ICU. These prediction scores, however, have a number of limitations, one of the most important being that they rely on patients’ data at admission, without re-evaluation during their stays. The prediction performances of these scores are therefore high regarding early complications, but show a decrease in their capacity to estimate the mortality risk in patients who have already spent several days or weeks in ICU [6].

To deal with this issue, other scoring systems have been developed to estimate the risk of complications throughout the stay, using updated collection of patients’ data. Regarding the risk of septic shock, one of the leading causes of death in ICU patients, longitudinal collection of data thus allowed to identify a “pre-shock” state during which the symptoms of the upcoming failure are not yet clinically visible [5]. Early management of this “pre-shock” state may allow to prevent the occurrence of a septic shock and to improve survival. Opportunities to predict or early identify the onset of complications therefore represent a major challenge in the management of ICU patients.

The current spread of healthcare data warehouses offers new opportunities to closely monitor the evolution of ICU inpatients and to develop prognostic scores relying on a wider range of data [7,8]. These databases enable the collection and centralization of detailed data throughout inpatients’ stays via demographic characteristics, physiological measures, diagnoses, laboratory analyses, medical imaging, medical notes, etc. ICUs are highly monitored environments and important data sources for these warehouses. Repeated collection of data allows to study the evolution of patients’ characteristics and to identify factors associated with the occurrence of worsening conditions, possibly leading to complications or death. Appropriate machine learning algorithms are required to address the massive amount of data available in these warehouses. Deep learning methods have been extensively studied in the recent years for their abilities to manage large amounts of data and specific architectures of deep learning networks, such as convolutional and recurrent neural networks, were dedicatedly developed to handle longitudinal data [7,9]. Such predictive modeling approaches may however present a limited interest when their use relies on a large amount of predictors, several of which may be unavailable in some patients.

In this study, we aimed to develop and validate models predicting ICU mortality for higher lengths of stay than those well evaluated by the existing scores. These models were built from ICU hospitalizations lasting more than 48 hours, using longitudinal healthcare data with missing values from electronic health records available in the freely accessible Medical Information Mart for Intensive Care (MIMIC-III) critical care database [10,11]. Different architectures of deep learning neural networks were evaluated in a context of missing values for some predictors and compared with predictive models based on penalized regression.

## 2. Material and methods

### 2.1. Data collection and preparation

All predictive models were trained from the MIMIC-III database (version: January 2020). This data warehouse is an open-access database that collected anonymized care data in 46,520 patients from 19 critical care units of the Beth Israel Deaconess Medical Center in Boston, USA, between 2001 and 2012. Only the first ICU stay of each patient in the MIMIC-III database was used. Patients aged under 15 or over 100 years were excluded, as well as those with missing information on vital status at hospital discharge and those with an ICU length of stay lower than 48 hours.

Data collected throughout patients stays were split into several time slots, during which information was summarized by a unique value per variable (Figure 1). Short-term evolution of all patients’ characteristics was accounted with the use of triplets for consecutive values over three time slots, these triplets being used as predictors for model development. Time slots durations of 6 and 12 hours were compared, with predictions still addressing mortality between 12 and 24 hours following the 3^rd^ predictive slot, in order to find a trade-off between the ability to capture short-term evolutions and the overall duration of data collection. An additional format with 6 consecutive 6-hour predictive time slots was also tested. In all analyses, the models aimed at predicting mortality after a 12-hour gap following the third predictive time slot. For instance, using 12-hour time slots following time t_0_, information collected over the 3 time slots between t_0_ and t_0_ + 36h were used to predict mortality between t_0_ + 48h and t_0_ + 60h. The 12-hour gap between t_0_ + 36h and t_0_ + 48h was considered clinically relevant as it is short enough to predict upcoming lethal complications, yet leaves some time for physicians to become aware of possibly undetected complications and modify diagnostic or therapeutic management if necessary. Unlike the current scores using admission data, these models therefore apply only in patients staying more than 48 hours in ICU.

**Figure 1.**
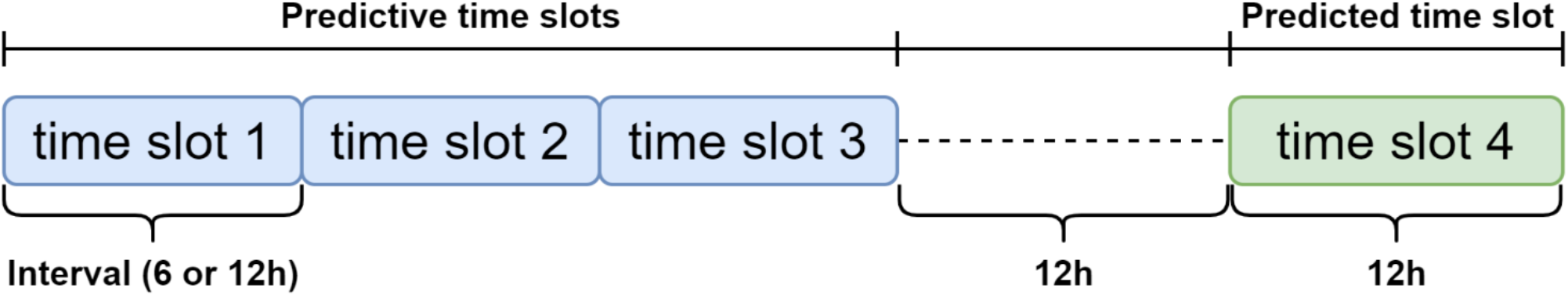
Time-slot formatting of data. For variables with values evolving within a predictive time slot, latest values were used. Durations of 6 and 12h were compared for predictive time slots.

The predicted endpoint was mortality in ICU, coded as a binary variable. Assuming that patients’ characteristics associated with mortality in ICU were mostly identified by previously published prognostic scores, we first developed models relying only on variables used in the APACHE II and SAPS II scores [3,4] to predict mortality, as well as the SOFA score [12] to predict the occurrence of organ failure. Nineteen predictors used in these scores were selected (Table 1), including medical history, vital signs, blood tests, as well as some administrative features such as previous hospitalization wards, which can provide information on the most common complications.

**Table 1.**
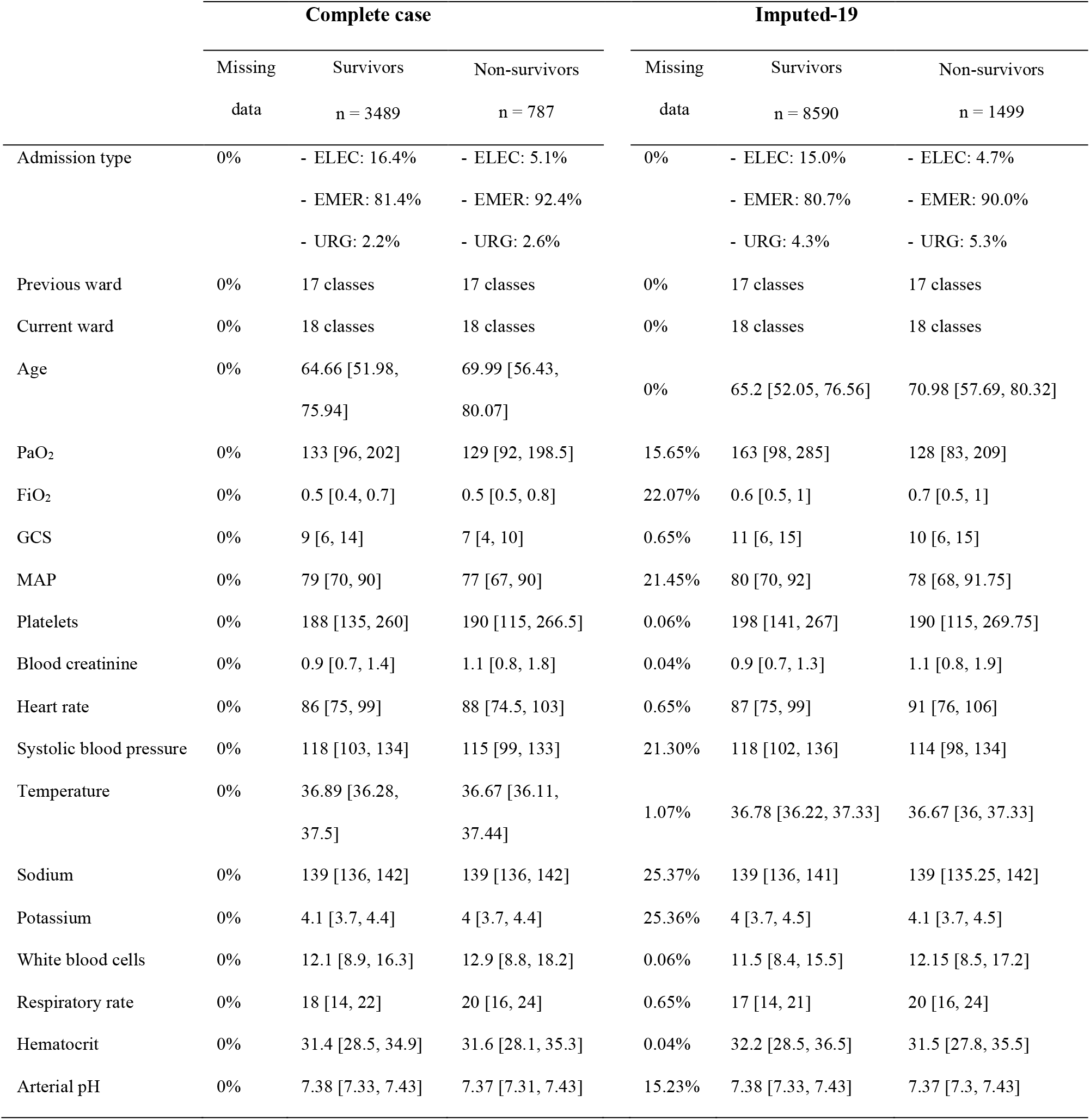
Characteristics of patients in the Complete case and Imputed-19 cohorts at admission. ELEC: elective; EMER: emergency; URG: urgent; PaO2: arterial partial pressure of oxygen; FiO_2_: fraction of inspired oxygen; GCS: Glasgow Coma Scale; MAP: mean arterial pressure

|  | Complete case |  |  | Imputed-19 |  |  |
| --- | --- | --- | --- | --- | --- | --- |
|  | Missing<br>data | Survivors<br>n = 3489 | Non-survivors<br>n = 787 | Missing<br>data | Survivors<br>n = 8590 | Non-survivors<br>n = 1499 |
| Admission type | 0% | - ELEC: 16.4%<br>- EMER: 81.4%<br>- URG: 2.2% | - ELEC: 5.1%<br>- EMER: 92.4%<br>- URG: 2.6% | 0% | - ELEC: 15.0%<br>- EMER: 80.7%<br>- URG: 4.3% | - ELEC: 4.7%<br>- EMER: 90.0%<br>- URG: 5.3% |
| Previous ward | 0% | 17 classes | 17 classes | 0% | 17 classes | 17 classes |
| Current ward | 0% | 18 classes | 18 classes | 0% | 18 classes | 18 classes |
| Age | 0% | 64.66 [51.98,<br>75.94] | 69.99 [56.43,<br>80.07] | 0% | 65.2 [52.05, 76.56] | 70.98 [57.69, 80.32] |
| PaO<sub>2</sub> | 0% | 133 [96, 202] | 129 [92, 198.5] | 15.65% | 163 [98, 285] | 128 [83, 209] |
| FiO<sub>2</sub> | 0% | 0.5 [0.4, 0.7] | 0.5 [0.5, 0.8] | 22.07% | 0.6 [0.5, 1] | 0.7 [0.5, 1] |
| GCS | 0% | 9 [6, 14] | 7 [4, 10] | 0.65% | 11 [6, 15] | 10 [6, 15] |
| MAP | 0% | 79 [70, 90] | 77 [67, 90] | 21.45% | 80 [70, 92] | 78 [68, 91.75] |
| Platelets | 0% | 188 [135, 260] | 190 [115, 266.5] | 0.06% | 198 [141, 267] | 190 [115, 269.75] |
| Blood creatinine | 0% | 0.9 [0.7, 1.4] | 1.1 [0.8, 1.8] | 0.04% | 0.9 [0.7, 1.3] | 1.1 [0.8, 1.9] |
| Heart rate | 0% | 86 [75, 99] | 88 [74.5, 103] | 0.65% | 87 [75, 99] | 91 [76, 106] |
| Systolic blood pressure | 0% | 118 [103, 134] | 115 [99, 133] | 21.30% | 118 [102, 136] | 114 [98, 134] |
| Temperature | 0% | 36.89 [36.28,<br>37.5] | 36.67 [36.11,<br>37.44] | 1.07% | 36.78 [36.22, 37.33] | 36.67 [36, 37.33] |
| Sodium | 0% | 139 [136, 142] | 139 [136, 142] | 25.37% | 139 [136, 141] | 139 [135.25, 142] |
| Potassium | 0% | 4.1 [3.7, 4.4] | 4 [3.7, 4.4] | 25.36% | 4 [3.7, 4.5] | 4.1 [3.7, 4.5] |
| White blood cells | 0% | 12.1 [8.9, 16.3] | 12.9 [8.8, 18.2] | 0.06% | 11.5 [8.4, 15.5] | 12.15 [8.5, 17.2] |
| Respiratory rate | 0% | 18 [14, 22] | 20 [16, 24] | 0.65% | 17 [14, 21] | 20 [16, 24] |
| Hematocrit | 0% | 31.4 [28.5, 34.9] | 31.6 [28.1, 35.3] | 0.04% | 32.2 [28.5, 36.5] | 31.5 [27.8, 35.5] |
| Arterial pH | 0% | 7.38 [7.33, 7.43] | 7.37 [7.31, 7.43] | 15.23% | 7.38 [7.33, 7.43] | 7.37 [7.3, 7.43] |

In order to assess the relevance of using longitudinal data, predictive models derived solely from admission data were built from the same dataset (thus only addressing patients alive and still in ICU 48h after admission to predict death anytime during their stays).

### 2.2. Missing data

Selected predictors were subject to missing values, to a large extent for some of them. Three approaches were compared to handle incomplete data. First, the analysis used the complete case cohort, by selecting only patients in whom all variables were available for the first time slot and “last observation carried forward” for the following slots. Second, missing values for the 19 selected predictors were imputed using multiple imputation by chained equations, with respect for the hierarchical structure of data (time slots within patients) [13,14], which allowed to keep additional patients in whom data was available for at least one of these 19 predictors. Third, the set of covariates used to predict ICU mortality was extended to a larger set of clinical and biological variables regardless of preexisting scores, and missing values for all variables were multiply imputed. This third approach allowed to consider a large extent of available predictors without limiting the sample size as would be required by the complete case analysis. A new set of 70 predictors was defined according to their availability among patients, which allowed to keep patients in whom data was available for at least one of these 70 predictors. These predictors were selected solely based on their availability, regardless of their expected clinical relevancy or collinearity (e.g., several predictors could describe the same measure performed by different devices, supplementary Table S1). Continuous predictors were log-transformed when required to improve normality. Parameters derived from multiple imputation were estimated with their standard errors through 10 imputed datasets and pooled using Rubin’s rule [15].

### 2.3. Neural Network architectures and statistical analyses

Four architectures of neural networks were set up to predict mortality in ICU inpatients: a fully connected neural network (FCN), a convolutional neural network (CNN) [16,17], a bidirectional long short-term memory (LSTM) recurrent neural network [18] and a CNN-LSTM network [19], which concatenated the information from the two previous networks.

The FCN used 6 dense layers with a decreasing number of neurons. The CNN used 3 convolutional layers, an average pooling layer and finally a fully connected layer allowing the classification done by the model. The LSTM network used a single LSTM layer with a fully connected output layer. Finally, the CNN-LSTM network combined the CNN and the LSTM networks (using the same hyper-parameters), with a concatenation layer and a dense layer as an output.

All neural network used Rectified Linear Unit (ReLU) activation functions in the hidden layers, and a dense output layer with two neurons (for two classes) and a sigmoid activation function. The parameters were optimized with a binary cross-entropy loss function, the Adam optimizer [20], and a learning rate of 0.001. Observations were weighted according to the outcome group they belonged to, in order to correct the imbalance between these groups [21].

Using the same data, these neural networks were compared with elastic net, a regularized logistic regression approach that combines the L1 and L2 penalties of the lasso and ridge methods to control multicollinearity, which commonly occurs in models with large numbers of predictors [22].

Internal validity and robustness of model predictions were assessed using multiple 5-fold cross-validation: patients were split into 5 subsets, 4 of which were used for model training and the 5^th^ for performances evaluation, this procedure being repeated so that all 5 subsets were used for evaluation. This 5-fold cross-validation was carried out 10 times with different partitions of the dataset. Models’ ability to discriminate patients at higher risk of death was evaluated using the average area under the ROC curve (AUC). Pairwise AUC comparisons were performed between models using linear mixed models with a random intercept for the cross-validation dataset partition. Fixed effects were tested directly for the complete case analyses and after pooling with Rubin’s rule for imputed datasets. Calibration was graphically assessed with calibration plots comparing observed and predicted probabilities [23], after rescaling of predictions according to the imbalance weights used for model training.

Neural networks were built using Keras version 2.3.1, the application programming interface of Tensorflow version 2.1.0. All other analyses were performed using R Statistical Software version 4.0.2 (Foundation for Statistical Computing, Vienna, Austria). All tests were two-tailed at the 0.05 significance threshold.

This study followed guidelines from the Transparent Reporting of a multivariable prediction model for Individual Prognosis Or Diagnosis (TRIPOD) statement [23]. The TRIPOD checklist is provided as supplementary Table S2.

## 3. Results

### 3.1. Selection of patients

After exclusion of patients aged < 15 or > 100 years, those with missing data on vital status and those staying < 48h in ICU, 17,373 patients with unique admission remained in the dataset. According to missing data management, three cohorts were defined. Patients with no missing data in the 19 initially selected predictors, at least for the first time slot, defined the “complete case” cohort (n = 4276 patients, 787 deaths). The “imputed-19” cohort included patients with data available for at least one of the 19 main predictors (n = 10,089 patients, 1499 deaths), whereas the “imputed-70” cohort did the same with the extended selection of 70 predictors (n = 16,532 patients, 2395 deaths). Models were derived from these two latter cohorts after multiple imputation. Figure 2 summarizes this selection process.

**Figure 2.**
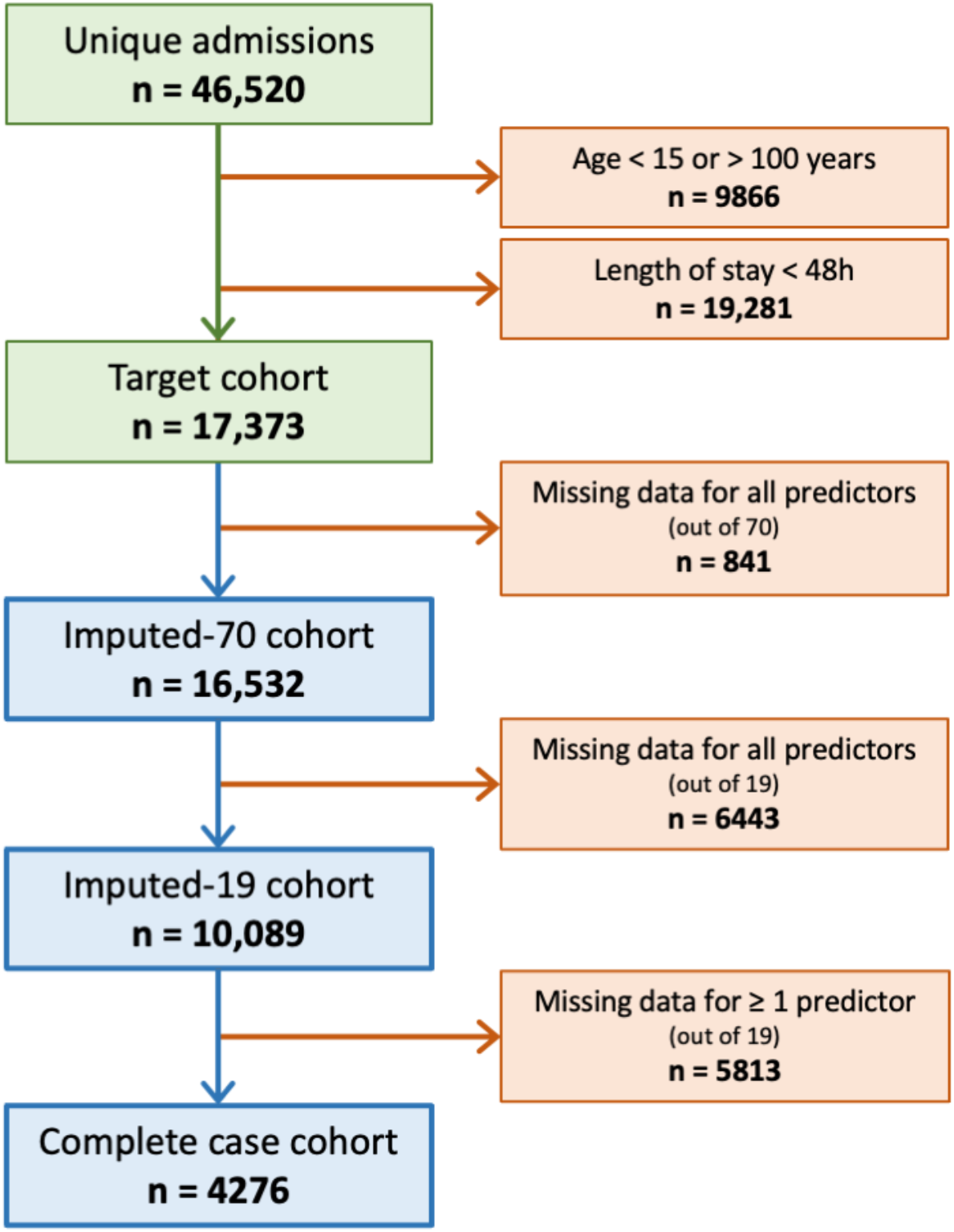
Flowchart for the definition of the three cohorts from the MIMIC-III database. Patients were selected according to age, length of stay ≥ 48 hour and available data among the selected predictors.

### 3.2. Models derived from data at admission

Using only admission data from the complete case cohort to predict death in ICU in patients still in ICU 48h after admission, the CNN showed the best performances (AUC = 0.742 ± 0.002, p < 0.001 compared with any other method). The elastic net ranked second (AUC = 0.709 ± 0.002), while the FCN, the LSTM and the CNN-LSTM all had AUC under 0.67 (Table 2).

**Table 2.** Performance of elastic net and neural networks to predict ICU mortality (AUC ± SE). Predictions based on admission data or longitudinal data with either 6-hour or 12-hour slots are compared for the complete case cohort only. Cohorts defined by missing data management (Complete case, Imputed-19 or Imputed-70 cohorts) are compared for predictions based on 12-hour slots only. AUC: Area under the ROC curve; SE: Standard error; FCN: Fully connected network; CNN: Convolutional neural network; LSTM: Long short-term memory. Imputed-19: Missing values imputed for the same 19 predictors as complete case analyses; Imputed-70: Missing values imputed for an additional set of 51 predictors.

|  | Complete case |  |  | Imputed-19 | Imputed-70 |
| --- | --- | --- | --- | --- | --- |
|  | n = 4276 |  |  | n = 10,089 | n = 16,532 |
|  | Admission data | 6-hour slots | 12-hour slots | 12-hour slots | 12-hour slots |
| **Elastic net** | 0.709 ± 0.002 | 0.769 ± 0.029 | 0.785 ± 0.002 | 0.753 ± 0.024 | 0.777 ± 0.003 |
| **FCN** | 0.663 ± 0.055 | 0.521 ± 0.037 | 0.634 ± 0.049 | 0.586 ± 0.056 | 0.542 ± 0.047 |
| **CNN** | 0.742 ± 0.002 | 0.778 ± 0.005 | 0.778 ± 0.006 | 0.751 ± 0.022 | 0.783 ± 0.003 |
| **LSTM** | 0.602 ± 0.027 | 0.751 ± 0.028 | 0.773 ± 0.016 | 0.764 ± 0.017 | 0.775 ± 0.019 |
| **CNN-LSTM** | 0.663 ± 0.028 | 0.780 ± 0.003 | 0.770 ± 0.009 | 0.772 ± 0.004 | 0.779 ± 0.018 |

### 3.3. Time slots duration

Now relying on longitudinal data, still with the complete case cohort, neural networks performances using time slots of 6- and 12-hours durations were compared. Fully connected networks showed poorer performances than the rest of the models for all time slots duration (p < 0.001 compared with any other method). Models with the best performances were elastic net for 12-hour slots (AUC = 0.789 ± 0.002) and CNN-LSTM for 6-hour slots (AUC = 0.780 ± 0.003), with similar AUC (p = 0.193). Except for the FCN that always showed poor performances, all methods using longitudinal data with either 6-or 12-hour slots outperformed the same methods using only admission data (p < 0.001 for all methods).

### 3.4. Missing data handling

Multiple imputation of missing values allowed to consider a larger set of predictors and to keep larger sample sizes than for complete case analyses. Table 2 summarizes the predictive performances for all cohorts with 12-hour time slots.

#### Imputed-19

Multiple imputation of the 19 previous predictors allowed to include nearly 2.5 times as many patients as in the complete case analysis, with similar or slightly weaker performances. The CNN-LSTM showed the best performances (AUC = 0.772 ± 0.004, p < 0.001 compared with any other method), close to the complete case analysis, followed by the LSTM (AUC = 0.764 ± 0.017), the elastic net model (AUC = 0.753 ± 0.024) and the CNN (AUC = 0.751 ± 0.022).

#### Imputed-70

Extending the set of predictors to 70 covariates allowed to include nearly 4 times as many patients as in the complete case analysis, with similar or slightly better performances except for the FCN. The CNN and the CNN-LSTM showed the best performances (AUC = 0.783 ± 0.003 and 0.779 ± 0.018, p = 0.095). The elastic net (AUC = 0.777 ± 0.003) and the LSTM (AUC = 0.775 ± 0.019) showed poorer performances (p < 0.001 compared with the CNN).

Figure 3 summarizes the discrimination and calibration performances of the compared modeling methods for all cohorts, except for the FCN which demonstrated poor performances in all analyses. ROC curves and calibration plots represent the average estimates over the 10 repeated 5-fold cross validation, and over the imputed datasets for the Imputed-19 and Imputed-70 cohorts.

**Figure 3.**
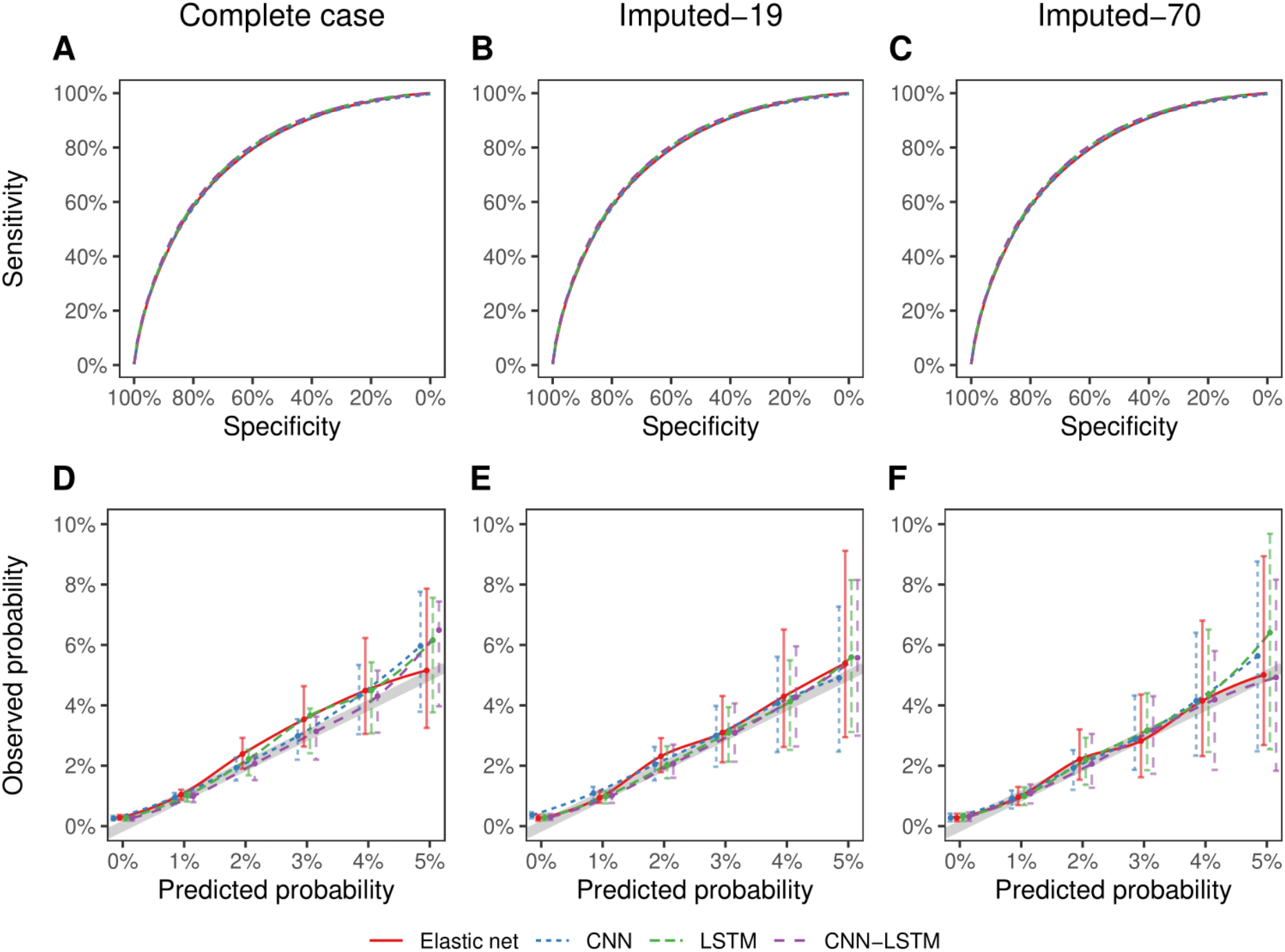
Predictive performances of the elastic net, CNN, LSTM and LSTM-CNN models. Discrimination is represented by the ROC curve (upper figures) and calibration by a smoothed calibration plot showing the observed probabilities (and 95% confidence intervals) according to predicted probabilities. The thick gray line shows values expected for a perfect calibration, with observed probabilities equal to predicted probabilities. All estimates are averaged over the 10 repeated 5-fold cross validation datasets, and over the imputed datasets for the Imputed-19 and Imputed-70 cohorts.

All models globally present a fairly satisfactory calibration.

## 4. Discussion

This study aimed to assess the relevancy of predictive models for mortality in intermediate or long-term ICU stays, relying on healthcare data iteratively collected throughout patients’ stays to reevaluate patients’ prognoses. Complementary to usual predictive scores for mortality occurring shortly after admission, such as SAPS-II or APACHE II, our analyses focused on patients staying at least 48 hours in ICU.

Mortality risk after 48 hours is estimated continuously using data collected shortly before. We nevertheless considered a minimal 12-hour gap between the end of data collection and death, so that the developed models do not identify premortem status but rather leave some time for the medical staff to handle the situation. Considering the massive amount of data possibly available in data warehouses, we focused on statistical approaches likely to integrate a large number of variables, such as deep neural networks and penalized regression models.

We found that the 6- and 12-hour formats provided similar performances. The 12-hour format, as it allows to update predictions twice a day at predetermined times or on a regular basis, probably corresponds to the most relevant time slot size.

Some of the models we developed show performances very close to the classical predictive scores of ICU mortality [4,12] or other ICU mortality prediction models based on neural networks [24], yet these latter models use admission data that are not updated throughout patients’ stays. Our first analyses confirmed that the integration of data collected during patients’ stays permitted to identify patients at higher risk of death better than when relying on baseline data only. Though unsurprising, this result highlights the need to develop and validate predictive scores that could more accurately evaluate patients’ prognoses after some time spent in ICU.

Missing data is an important issue in clinical studies [25,26], causing a several problems for complete case analysis: the decreased sample size hampers the training of models, while the exclusion of patients possibly yields selection biases that affects models’ performances and proscribes predictions in patients with incomplete data. Using data previously identified as predictive of ICU mortality, our complete case analysis showed satisfactory results, with AUC between 0.77 and 0.79 for both penalized regression and convolutional neural networks. However, including patients with data available for all predictors implied to select a subsample of only 4276 out of the 17,373 in the target cohort (25%), which suggests both a possible selection bias and the inability of our models to infer a mortality risk for patients in whom some of these predictors would be missing.

Multiple imputation by chained equations, using available information for a given patient and the associations between variables derived from the whole sample [27], appeared as a promising option to address these issues. A first attempt to impute data for these 19 predictors (Imputed-19 cohort) allowed to include a larger sample size (10,089 patients, 58% of the target cohort) without degrading predictive performances. More interestingly, data imputation considering a larger set of potential predictors (Imputed-70 cohort) allowed to include an even larger sample size (16,532 patients, 95% of the target cohort) with slightly better performances than for the complete case cohort. The excluded patients are those in whom no data was available at admission and it is therefore difficult to figure out how they differ from included patients. For similar reasons, data imputation relies on a hypothesis of “missing completely at random” (MCAR) or “missing at random” (MAR) mechanisms and we cannot rule out a “missing not at random” (MNAR) mechanism (the probability of missing values depends on unobserved characteristics). In such a context, our models would yield biased estimates in patients with data missing for specific predictors. However, our cross-validation procedure used to estimate models’ performances captures the inaccuracy that could result from the missing data pattern and reported results already integrate this possible source of error. This robustness to missing data imputation is insightful as it allows to consider that predictive models might be developed in ICUs admitting patients with specific conditions and provide prognosis predictions for all patients, with a higher precision as available information accumulates.

Though deep neural networks are increasingly popular to handle massive data, they did not outperform more conventional penalized regressions in our study. An explanation might be that available data did not take full advantage of the time slots format [28]: though some predictors, such as vital signs or blood tests, were frequently updated, medical conditions likely to dramatically impact prognosis such as the occurrence of shock or organ failure were collected retrospectively but not on time to be used as a predictor. This limitation is due to the nature of the MIMIC-III database and may be present in other healthcare data warehouses, yet we assume that a timely collection of medical diagnoses and relevant symptoms may be insightful to enhance predictive performances.

Our study has several strengths, including a novel approach to integrate updated information on patients’ characteristics to estimate their prognosis more accurately and the additional opportunity to use this information even when data is partially missing. The 12-hour gap between collection of predictors and occurrence of the predicted event also appears as clinically relevant as it allows the medical staff to take preventive measures whenever possible. Depending on the specificities of each ICU, similar predictive models could be developed for other outcomes than mortality, e.g., the occurrence of shock, organ failure or multiple organ dysfunction.

Several limitations must also be noticed. First, this study must be seen as a “proof of concept” for a novel predictive modelling framework but we do not expect that inferring our models’ parameters to other ICUs with specific patients and data collections might yield meaningful predictions. We nevertheless assume that using the same modelling approaches in a new setting may produce models with similar performances. Additionally, contrary to exponentiated regression coefficients of elastic net models that can be directly interpreted as odds ratios for the considered predictors, the “black box” nature of neural networks does not allow to easily identify specific predictors associated with a higher risk of mortality. These models must therefore be seen as global “alert systems” rather than as a tool likely to identify specific complications. Finally, a technical limitation relies on the possibility of data collection and automated analysis of healthcare data almost in real time. Though very few ICUs might present this ability nowadays, the current development of healthcare data warehouses worldwide may enhance feasibility.

## Data Availability

All data used in the present study are publicly available: https://archive.physionet.org/physiobank/database/mimic3cdb

