## Supplemental material for "Updating mortality risk estimation in intensive care units from high-dimensional electronic health records with incomplete data"

### Supplementary material

- **Supplementary table S1.** Characteristics of patients in the Imputed-70 cohort at admission.
- **Supplementary table S2.** TRIPOD checklist for prediction model development and validation

| | Missing data | Survivors<br>n = 14,137 | Non-survivors<br>n = 2,395 |
|---|---|---|---|
| Male gender | 55.7% | 57.1% | 53.8% |
| Admission type | 0% | - ELEC: 2063 (14.6%) | - ELEC: 111 (4.6%) |
| | | - EMER: 11624 (82.2%) | - EMER: 2196 (91.7%) |
| | | - URG: 450 (3.2%) | - URG: 88 (3.7%) |
| Previous ward | 0% | 17 classes | 17 classes |
| Current ward | 0% | 18 classes | 18 classes |
| Marital status | 60.5% | - single: 1600 (28.3%) | - single: 235 (26.8%) |
| | | - married: 2948 (52.1%) | - married: 443 (52.1%) |
| | | - divorced: 409(7.2%) | - divorced: 63 (7.2%) |
| | | - widowed: 695 (12.3%) | - widowed: 135 (15.4%) |
| | | - life partner: 2 (0.04%) | - life partner: 0 (0%) |
| History of falling within 3 months | 60.9% | 1394 (25.1%) | 303 (33.3%) |
| Secondary diagnosis | 60.9% | 5049 (90.9%) | 852 (93.7%) |
| Ambulatory aid | 60.9% | - bed rest: 5492 (98.9%) | - bed rest: 908 (99.9%) |
| | | - walker: 51 (0.9%) | - walker: 1 (0.1%) |
| | | - furniture: 10 (0.2%) | - furniture: 0 (0%) |
| IV saline lock | 60.9% | 5324 (95.9%) | 880 (96.8%) |
| Gait transferring | 60.9% | - bed rest: 4141 (74.5%) | - bed rest: 881 (96.9%) |
| | | - weak: 1176 (21.2%) | - weak: 12(1.3%) |
| | | - impaired: 237(4.3%) | - impaired: 16 (1.8%) |
| Mental status | 60.9% | - oriented to own ability: 3355 (60.4%) | - oriented to own ability: 101(11.1%) |
| | | - forgets limitations: 2198 (39.6%) | - forgets limitations: 808 (88.9%) |
| Age | 0% | 64.86 [52.18, 76.49] | 70.93 [58.42, 80.25] |
| GCS (total) | 38.92% | 11 [6, 15] | 10 [6, 15] |
| GCS (eye opening) 1 | 38.92% | 3 [1, 4] | 3 [1, 4] |
| GCS (eye opening) 2 | 60.89% | 4 [1, 4] | 3 [1, 4] |
| GCS (verbal response) 1 | 60.89% | 4 [1, 5] | 1 [1, 5] |
| GCS (verbal response) 2 | 38.92% | 1 [1, 5] | 1 [1, 5] |
| GCS (motor response) 1 | 60.89% | 6 [4, 6] | 6 [4, 6] |
| GCS (motor response) 2 | 38.92% | 6 [4, 6] | 5 [4, 6] |
| Pain | 61.61% | 0 [0, 3] | 0 [0, 0] |
| Braden score | 39.00% | 14 [12, 16] | 13 [12, 15] |
| Braden sensory perception | 60.92% | 3 [2, 4] | 3 [2, 3] |
| Braden moisture | 60.92% | 4 [3, 4] | 4 [3, 4] |
| Braden activity | 60.92% | 1 [1, 1] | 1 [1, 1] |
| Braden mobility | 60.92% | 3 [2, 3] | 2 [2, 3] |
| Braden nutrition | 60.92% | 2 [2, 3] | 2 [2, 2] |
| Braden friction shear | 60.92% | 2 [2, 3] | 2 [2, 2] |
| Systolic BP 1 | 61.47% | 113 [101, 122] | 109 [96, 120] |
| Systolic BP 1 | 52.05% | 119 [104, 136] | 116 [99, 135] |
| Systolic BP 1 | 40.04% | 116 [101, 133] | 112 [98, 130] |
| Diastolic BP 1 | 60.99% | 63 [54, 75] | 62 [51, 74] |

| | | | |
|---|---|---|---|
| Diastolic BP 2 | 52.05% | 61 [53, 70] | 59 [50, 69] |
| Diastolic BP 3 | 40.05% | 57 [48, 68] | 55 [46, 66] |
| Mean BP 1 | 60.98% | 76 [66, 88] | 73 [63, 87] |
| Mean bp 2 | 40.06% | 76 [66, 87] | 73.67 [63, 85] |
| Mean BP 3 | 52.01% | 80 [71, 92] | 78 [68, 91] |
| Glucose 1 | 38.22% | 2.13 [2.04, 2.23] | 2.15 [2.05, 2.26] |
| Glucose 2 | 55.75% | 2.11 [2.03, 2.2] | 2.13 [2.03, 2.24] |
| Glucose 3 | 48.09% | 2.13 [2.05, 2.23] | 2.16 [2.06, 2.27] |
| Heart rate 1 | 38.92% | 87 [75, 99] | 91 [76, 106] |
| Heart rate 2 | 60.80% | 86 [74, 100] | 89 [76.75, 105] |
| Respiratory rate 1 | 60.82% | 18 [15, 22] | 20 [16, 25] |
| Respiratory rate 2 | 38.92% | 17 [14, 21] | 20 [16, 24] |
| Potassium 1 | 38.22% | 4 [3.7, 4.5] | 4.1 [3.7, 4.6] |
| Potassium 2 | 55.75% | 4.1 [3.7, 4.5] | 4.1 [3.7, 4.6] |
| $HCO_3^-$ | 0.09% | 23 [21, 26] | 22 [19, 26] |
| $FiO_2$ | 52.26% | 0.6 [0.5, 1] | 0.7 [0.5, 1] |
| $O_2$ saturation 1 | 60.82% | 98 [96, 100] | 98 [95, 100] |
| $O_2$ saturation 2 | 38.92% | 99 [97, 100] | 98 [96, 100] |
| $O_2$ flow | 44.56% | 5 [3, 12] | 5 [3, 12] |
| Arterial base excess | 68.89% | 4 [3, 5] | 4 [3, 5] |
| Arterial $CO_2$ | 48.17% | 25 [22, 28] | 23 [19, 28] |
| Arterial $PaCO_2$ | 48.17% | 40 [35, 46] | 38 [33, 46] |
| Arterial $PaO_2$ | 48.16% | 163 [98, 285] | 128 [83, 209] |
| Red blood cells | 38.31% | 3.57 [3.15, 4.05] | 3.48 [3.04, 3.95] |
| White blood cells | 0.27% | 1.05 [0.91, 1.18] | 1.08 [0.92, 1.23] |
| Hemoglobin | 0.07% | 10.8 [9.5, 12.3] | 10.6 [9.3, 12] |
| Hematocrit | 0.05% | 32.2 [28.4, 36.5] | 31.6 [27.7, 35.9] |
| Platelets | 0.07% | 2.3 [2.16, 2.43] | 2.28 [2.07, 2.44] |
| BUN | 0.04% | 1.26 [1.11, 1.45] | 1.41 [1.23, 1.64] |
| Calcium | 2.03% | 8.3 [7.7, 8.8] | 8.2 [7.6, 8.7] |
| Chloride | 0.06% | 106 [102, 109.25] | 105 [100, 109] |
| Creatinine | 0.13% | -0.05 [-0.15, 0.11] | 0.04 [-0.1, 0.28] |
| Magnesium | 0.19% | 1.9 [1.7, 2.2] | 1.9 [1.7, 2.2] |
| Sodium | 0.05% | 139 [136, 141] | 139 [135, 142] |
| Phosphorous | 1.86% | 0.54 [0.45, 0.62] | 0.57 [0.46, 0.67] |
| Arterial pH | 47.99% | 7.38 [7.33, 7.43] | 7.37 [7.3, 7.43] |
| Temperature | 0.47% | 36.78 [36.22, 37.33] | 36.61 [36, 37.22] |
| Weight | 4.48% | 80 [67.2, 94.6] | 75.5 [63, 90.4] |
| Previous weight | 49.48% | 82.1 [69, 97.5] | 78 [65.2, 93.9] |

**Supplementary table S1. Characteristics of patients in the Imputed-70 cohort at admission.**
$PaO_2$: Arterial partial pressure of oxygen; $FiO_2$: Fraction of inspired oxygen; GCS: Glasgow Coma Scale; BP: Blood pressure; $PaO_2$: Partial pressure of oxygen; $PaCO_2$: Partial pressure of carbon dioxide; BUN: Blood Urea Nitrogen

| Section/Topic | Item | Checklist Item | Page |
|---|---|---|---|
| **Title and abstract** | | | |
| Title | 1 | Identify the study as developing and/or validating a multivariable prediction model, the target population, and the outcome to be predicted. | 1 |
| Abstract | 2 | Provide a summary of objectives, study design, setting, participants, sample size, predictors, outcome, statistical analysis, results, and conclusions. | 1-2 |
| **Introduction** | | | |
| Background and objectives | 3a | Explain the medical context (including whether diagnostic or prognostic) and rationale for developing or validating the multivariable prediction model, including references to existing models. | 3-4 |
| | 3b | Specify the objectives, including whether the study describes the development or validation of the model or both. | 4 |
| **Methods** | | | |
| Source of data | 4a | Describe the study design or source of data (e.g., randomized trial, cohort, or registry data), separately for the development and validation data sets, if applicable. | 5 |
| | 4b | Specify the key study dates, including start of accrual; end of accrual; and, if applicable, end of follow-up. | - |
| Participants | 5a | Specify key elements of the study setting (e.g., primary care, secondary care, general population) including number and location of centres. | 5 |
| | 5b | Describe eligibility criteria for participants. | 5 |
| | 5c | Give details of treatments received, if relevant. | - |
| Outcome | 6a | Clearly define the outcome that is predicted by the prediction model, including how and when assessed. | 6 |
| | 6b | Report any actions to blind assessment of the outcome to be predicted. | - |
| Predictors | 7a | Clearly define all predictors used in developing or validating the multivariable prediction model, including how and when they were measured. | 5 - 7 |
| | 7b | Report any actions to blind assessment of predictors for the outcome and other predictors. | - |
| Sample size | 8 | Explain how the study size was arrived at. | 11 |
| Missing data | 9 | Describe how missing data were handled (e.g., complete-case analysis, single imputation, multiple imputation) with details of any imputation method. | 8 |
| Statistical analysis methods | 10a | Describe how predictors were handled in the analyses. | 5 - 6 |
| | 10b | Specify type of model, all model-building procedures (including any predictor selection), and method for internal validation. | 9 - 10 |
| | 10d | Specify all measures used to assess model performance and, if relevant, to compare multiple models. | 10 |
| Risk groups | 11 | Provide details on how risk groups were created, if done. | - |
| **Results** | | | |
| Participants | 13a | Describe the flow of participants through the study, including the number of participants with and without the outcome and, if applicable, a summary of the follow-up time. A diagram may be helpful. | 10 - 11 |
| | 13b | Describe the characteristics of the participants (basic demographics, clinical features, available predictors), including the number of participants with missing data for predictors and outcome. | 7 |
| Model development | 14a | Specify the number of participants and outcome events in each analysis. | 11 |
| | 14b | If done, report the unadjusted association between each candidate predictor and outcome. | - |
| Model specification | 15a | Present the full prediction model to allow predictions for individuals (i.e., all regression coefficients, and model intercept or baseline survival at a given time point). | - |
| | 15b | Explain how to the use the prediction model. | - |
| Model performance | 16 | Report performance measures (with CIs) for the prediction model. | 12 - 15 |
| **Discussion** | | | |
| Limitations | 18 | Discuss any limitations of the study (such as nonrepresentative sample, few events per predictor, missing data). | 18 - 19 |
| Interpretation | 19b | Give an overall interpretation of the results, considering objectives, limitations, and results from similar studies, and other relevant evidence. | 16 - 19 |
| Implications | 20 | Discuss the potential clinical use of the model and implications for future research. | 19 |
| **Other information** | | | |
| Supplementary information | 21 | Provide information about the availability of supplementary resources, such as study protocol, Web calculator, and data sets. | - |
| Funding | 22 | Give the source of funding and the role of the funders for the present study. | - |

**Supplementary table S2. TRIPOD checklist of items of interest in study developing a multivariable prediction model for prognosis.**
